# Leaders’ gender and the fight against COVID-19: investigation, replication, and a possible explanation

**DOI:** 10.1101/2021.10.12.21264903

**Authors:** Yossi Maaravi, Tamar Gur, Yossi Gur Arie

## Abstract

**Objectives:** To investigate the alleged relationship between leaders’ gender and COVID-19 related cases and deaths in different countries across the globe.

**Study design:** The relationship between leaders’ gender and percent of women in parliament to COVID-19 cases and death per million was investigated in three time points (10 months) across 180 countries, controlling for possible covariates.

**Methods:** Three different types of analyses were run: (1) Six basic t-tests; (2) Two repeated-measure ANOVA tests analyzing change over time; (3) Six stepwise regression analyses for both leaders’ gender and the percentage of women in parliament.

**Results:** Our findings suggest that, contrary to some research and popular media headlines, and in-line with recent academic research, leadership gender is not a significant factor in explaining the variation between countries in the fight against the COVID-19 pandemic.

**Conclusions:** We suggest that this alleged relationship may stem from an illusory correlation. We argue that the uncertainty, stress, and anxiety that prevail in these times of global pandemic may be the breeding ground for judgmental biases of this sort.

## Introduction

In the past year, COVID-19 has spread globally, killing almost three million people. To assist governments in fighting the pandemic, scientists have been studying various factors for contagion and prevention. Despite these unprecedented scientific efforts, one crucial question remains unanswered: why did COVID-19 hit some countries harder than others? Besides the “ immediate suspects” such as population age or health system quality, scientists have started investigating social factors such as countries’ individualism vs. collectivism^1^ or gender differences in political leadership^2^. Leadership gender draws special attention as it might have significant political and societal consequences. But, while prominent media outlets such as Forbes^3^ celebrated female leaders’ success, scientists have reached mixed results^2,4^.

The belief in this relationship may be rooted in scientific research to some extent^5^, but the most comprehensive research thus far has not supported it^2^. We argue that this belief may stem from gender stereotypes, which play a crucial role in human judgments, especially in times of uncertainty^6^. In the case of gender, people tend to ascribe different traits, behaviors, and beliefs to women and men. For example, whereas the “ typical” woman is judged to be warm, expressive, emphasizing communication, gentle, emotional, and passive, the “ typical” man is seen as tough, aggressive, emphasizing action, assertive, rational, and active^6^. Such stereotypes are also prevalent in politics and may influence citizens’ judgments of political candidates and leaders and voters’ voting patterns^7^.

Illusory correlation is a well-established judgmental bias that is considered a possible cognitive basis for the phenomenon of stereotype formation^8^. Decision-makers are often insensitive to the actual relation between variables or events and consequently perceive erroneous covariation or correlation, which are more prevalent under uncertainty. More relevant to the current article is the tendency to form illusory correlations when facing a rare event or occurrence^8^. For example, people see an illusory correlation between AIDS (a rare event) and gay males (a minority group). However, the incidence of this disease is higher among other groups, such as heterosexual women^9^. Analogically, it is possible that when trying to explain the rare success of a specific country in combating the pandemic (e.g., in New Zealand), people have mistakenly attributed it to its female leader (a minority group).

This perceived association was further cemented when New Zealand, led by Prime Minister Jacinda Ardern, became the first country to declare itself a COVID-19 virus-free nation. But, is this relationship a scientific conclusion or rather an illusory correlation? Recent scientific research^2^ has focused on the first stage of the pandemic (30, 60, 90, and 120 days after the outbreak) and suggested the latter. This short report expands this research line by investigating the relationship between leaders’ gender to COVID-19 cases and death in three time points (10 months) across 180 countries.

### The current research

The data used in this analysis were retrieved from Windsor et al.^2^ and Our World in Data^10^. While Windsor et al. ^2^ only studied the first phase of the pandemic (30, 60, 90, and 120 days after the outbreak), we investigated three time points that spanned ten months: May 1 2020, Sept 1, and Mar 1 2021. The data included a total of 180 countries. As detailed below, we analyzed the relationship of both leaders’ gender and for the percentage of women in parliament to COVID-19 cases and COVID-19 related deaths per million for each country, in each of the three time points. We also included the following covariates used in Windsor et al.^2^: Freedom House status (i.e., people’s access to political rights and civil liberties, coded as; Not Free, Partly Free; Free); GDP per capita; the percentage of the population over 65; land area; land borders and life expectancy.

## Methods

Altogether, we ran three different *types* of analyses to assess the relationship between leaders’ gender and COVID-19 cases and deaths: (1) Six basic t-tests - comparing cases and deaths between countries led by women and countries led by men in each of the three time points (Table 1); (2) Two repeated-measure ANOVA tests – analyzing the *change* in COVID-19 cases and deaths by leaders’ gender through time (Table 1); (3) Six stepwise regression analyses - examining the association between leaders gender and the percentage of women in parliament with the number of cases and deaths for each of the three time points (See supplementary materials for the complete analyses).

**Table 1.** T-test results comparing the number of reported cases and deaths for each of the timepoints in countries led by men and women.

|  | *t (df)* | *p* | *N* |
| --- | --- | --- | --- |
| Cases May 1<sup>st</sup> (20) | -1.23 (177) | .222 | 179 |
| Cases Sept' 1<sup>st</sup> (20) | .54 (178) | .593 | 180 |
| Cases Mar' 1<sup>st</sup> (21) | .27 (178) | .790 | 180 |
| Cases: Time | 28.423 (1.05, 186.38) | <.001 |  |
| Time * gender | .11 (1.05, 186.38) | .756 |  |
| Deaths May 1<sup>st</sup> (20) | -.77 (153) | .440 | 155 |
| Deaths Sept' 1<sup>st</sup> (20) | -.46 (166) | .646 | 168 |
| Deaths Mar' 1<sup>st</sup> (21) | .19 (173) | .850 | 175 |
| Deaths: Time | 27.93 (1.09, 166.43) | <.001 |  |
| Time * gender | -.63 (1.09, 166.43) | .442 |  |

Note: A negative sign indicates more cases/deaths per capita in men-led countries, while a positive sign means more cases/deaths per capita in women-led countries.

## Results

As shown in Table 1, there were no statistically significant differences between countries led by men versus those led by women for neither COVID -19 cases nor in fatalities in any of the three time points. Additionally, while the repeated-measure ANOVA tests revealed an expected significant effect of time (as the number of cases and fatalities naturally increased over time), we found no significant long-term difference between countries led by men compared with countries led by women. Next, we conducted six stepwise regression analyses to examine the association between leaders’ gender, the percentage of women in parliament as another indicator of leaderships gender, and COVID-19 cases and deaths per million for each of the three time points, controlling for the six covariates detailed above. Surprisingly, and in contrast to the popular media, statistically significant *positive* relationships were found between the percentage of women in parliament and both cases and deaths. As can be seen in the supplementary materials, this relationship appeared for COVID-19 cases in two of the time points and the number of deaths in all three time points.

We believe that this relationship does not suggest any causality but represents a statistical artefact that stems from other factors. Specifically, we suggest that countries where many women serve in the parliament share other characteristics that may explain both the spread of the pandemic and women’s participation in the political arena. One such factor that has been recently connected to the spread of the pandemic is the level of countries’ individualism (as opposed to collectivism)^1^. Indeed, we found a significant positive relationship between countries’ individualism and the percentage of women in the parliament (*r* = .28, *p* = .022). We also found a significant positive relationship between countries’ indulgence levels (e.g., allowing gratification of personal needs and reduced social regulation) and the percentage of women in the parliament (*r* = .22, *p* = .035). When holding both individualism and indulgence, these relationships disappeared. The number of cases was not associated with the percentage of women in parliament. The number of deaths was related to the percentage of women in the parliament only in the first data point.

## Conclusion

Countries led by women leaders did not seem to fare better or worse in the COVID-19 pandemic than countries led by men. This pattern did not change over time as the number of COVID-19 cases and deaths continued to grow. This may be a real-world demonstration of illusory correlation and a possible formation of a new gender stereotype. A few female leaders may indeed deserve compliments for their response to the crisis (e.g., New Zealand Prime Minister, Mrs. Jacinda Ardern). But their success may not necessarily be related to their gender but rather to their professional merits.

Moreover, gender equality is an important social issue with far-reaching consequences. Thus, emphasizing the role of leaders’ gender in fighting COVID-19 without providing scientific support may lead to the opposite results. Indeed, such popular theories may cause a “ boomerang” effect, where opponents of gender equality automatically label similar news in the future as “ fake news.”

## Data Availability

Data availability statement: The data that support the findings of this study are openly available at https://osf.io/g3n4v/?view_only=915933f0c4e745fca3bf66549d6fa23c

## Ethical approval

We got our institution’s ethical committee approval for this research.

## Supplementary Materials

The data that support the findings of this study are openly available at **https://osf.io/g3n4v/?view_only=915933f0c4e745fca3bf66549d6fa23c**

